# COVID-19 redux: clinical, virologic, and immunologic evaluation of clinical rebound after nirmatrelvir/ritonavir

**DOI:** 10.1101/2022.06.16.22276392

**Authors:** Brian P. Epling, Joseph M. Rocco, Kristin L. Boswell, Elizabeth Laidlaw, Frances Galindo, Anela Kellogg, Sanchita Das, Allison Roder, Elodie Ghedin, Allie Kreitman, Robin L. Dewar, Sophie E. M. Kelly, Heather Kalish, Tauseef Rehman, Jeroen Highbarger, Adam Rupert, Gregory Kocher, Michael R. Holbrook, Andrea Lisco, Maura Manion, Richard A. Koup, Irini Sereti

**Author notes:** Corresponding Author Irini Sereti M.D., Chief, HIV Pathogenesis Section, Laboratory of Immunoregulation, National Institute of Allergy & Infectious Diseases, National Institutes of Health, BG 10 RM 11B17 MSC 1876, 10 Center Dr, Bethesda, MD 20892. Equal contribution, names listed alphabetically (Brian P. Epling, Joseph M. Rocco).

## Abstract

Clinical rebound of COVID-19 after nirmatrelvir/ritonavir treatment has been reported. We performed clinical, virologic, and immune measurements in seven patients with symptomatic rebound, six after nirmatrelvir/ritonavir treatment and one without previous treatment. There was no evidence of severe disease or impaired antibody and T-cell responses in people with rebound symptoms.

## Introduction

Nirmatrelvir-ritonavir (NMV-r) has been granted an Emergency Use Authorization for treatment of early mild-moderate COVID-19 after demonstrating an 89% relative risk reduction of hospitalization or death in unvaccinated patients at high risk for severe disease, with an associated decrease in nasopharyngeal viral load at day-five.^1,2^ Some patients demonstrate a rise in viral load between days 10-14,^1^ and clinical rebound after completing NMV-r has now been reported.^3^ The etiology of this rebound remains unknown, and immune evasion because of early viral suppression has been hypothesized.^4^ Additionally, the risk of severe disease, viral transmissibility, and potential for NMV-r resistance remains unclear. To address these questions, we performed detailed virologic and immunologic evaluations of seven patients with rebound COVID-19.

## Methods

Detailed methods are provided in supplement. All patients were evaluated after written informed consent and enrollment in clinical protocol NCT04401436. Soluble biomarkers were measured using enzyme-linked immunosorbent assay (ELISA) or electrochemiluminescence assays. Viral isolation on Vero E6 cells and targeted viral sequencing were performed on SARS-CoV-2 positive nasal swab samples, as previously described.^5,6^ Serum SARS-CoV-2 nucleocapsid protein was measured using a single-molecule immunobead assay.^7^ ELISA was performed for SARS-CoV-2 anti-spike (anti-S) and anti-receptor binding domain (anti-RBD) IgG, IgA, and IgM and for IgG and IgM anti-nucleocapsid antibodies using a previously validated technique^8^. The GenScript cPass assay, a surrogate viral neutralization test (sVNT)^9^, was done to detect neutralizing antibodies against wild-type and Omicron spike protein. T-cell stimulation assays using peptide pools corresponding to SARS-CoV-2 spike, nucleocapsid, and membrane proteins were performed.

## Results

Seven patients with rebound COVID-19 symptoms (six following NMV-r, one without treatment) were seen, two with samples also collected during their acute presentation. Five COVID-19 (Omicron)-infected patients were included as controls. Based on time from symptom onset to sample collection, patients were subdivided into an acute (<4-days, n=6) and late group (>11-days, n=2, including the patient with rebound without prior NMV-r) that were compared to the rebound patients who received NMV-r (n=6) (Table-1, Table-S1). All participants were vaccinated/boosted, and none received CYP3A4 inducers prior to NMV-r.

**Table 1:** Patient Characteristics

|  | **Acute<sup>a</sup>**<br>**(N=6)** | **Rebound after NMV-r**<br>**(N=6)** | **Late<sup>b</sup>**<br>**(N=2)** |
| --- | --- | --- | --- |
| **Gender** |  |  |  |
| Male | 2 (33%) | 3 (50%) | 1 (50%) |
| Female | 4 (67%) | 3 (50%) | 1 (50%) |
| **Age (years)** |  |  |  |
| Median [Min, Max] | 49.5 [33, 65] | 42.5 [33, 74] | 38.5 [23, 54] |
| **Comorbidities** |  |  |  |
| None | 1 (17%) | 0 (0%) | 2 (100%) |
| Pulmonary | 1 (17%) | 2 (33%) | 0 (0%) |
| Immunocompromised<sup>c</sup> | 2 (33%) | 2 (33%) | 0 (0%) |
| Cardiac | 0 (0%) | 2 (33%) | 0 (0%) |
| Other | 2 (33%) | 0 (0%) | 0 (0%) |
| **Days from initial symptom onset to visit** |  |  |  |
| Median [Min, Max] | 3 [2, 4] | 16 [11, 17] | 11 [8, 14] |
| **Initial symptoms** |  |  |  |
| Upper respiratory only | 2 (33%) | 2 (33%) | 0 (0%) |
| Upper and lower respiratory symptoms | 2 (33%) | 1 (17%) | 1 (50%) |
| Upper respiratory and constitutional | 1 (17%) | 3 (50%) | 1 (50%) |
| Upper respiratory, lower respiratory, and constitutional | 1 (17%) | 0 (0%) | 0 (0%) |
| **Received NMV-r** |  |  |  |
| NMV-r Recipients | 4 (67%) | 6 (100%) | 0 (0%) |
| NMV-r Nonrecipients | 2 (33%) | 0 (0%) | 2 (100%) |
| **Day of illness NMV-r started** |  |  |  |
| Median [Min, Max] | 2.50 [2, 3] | 2.50 [1, 4] | NA |
| **Day of illness symptoms returned** |  |  |  |
| Median [Min, Max] | 13 [11, 15] | 12.5 [11, 15]<sup>d</sup> | 9 [rebound patient only] |
| **Day symptoms returned after completing NMV-r** |  |  |  |
| Median [Min, Max] | 6 [4, 8] | 6.50 [3, 9] | NA |

<sup>a</sup> Two patients were seen during both their initial acute episode as well as during rebound symptoms. Their first timepoint was included in the “acute” group, and their second timepoint in the “NMV-r rebound” group. They both
had symptom resolution and negative antigen tests after completing NMV-r with symptom return and positive antigen tests at eight and five days after completing NMV-r, respectively. See supplemental table 3 for further details (patient 1 and patient 2).
<sup>b</sup> The NMV-r nonrecipient rebound patient presented 14 days after symptom onset and was included alongside a control who had presented 8 days after symptom onset in the “late” group.
<sup>c</sup> Immunocompromising conditions across the entire cohort include multiple sclerosis, idiopathic thrombocytopenic purpura, ankylosing spondylitis, and primary biliary cirrhosis
<sup>d</sup> Four rebound patients repeated rapid antigen tests after initial symptom resolution. Three became negative and the fourth became weakly positive. All four became positive again when rebound symptoms returned.

NMV-r was started 1-4 days after initial symptom onset. All rebound patients experienced significant symptomatic improvement prior to worsening. Median time to symptom recurrence was 12.5-days after initial symptom onset and 6.5-days after completing NMV-r. On rebound, five patients reported milder, one worse and one similar symptom severity from initial illness (Table-S2). No additional pathogens were detected by BioFire® FilmArray® Respiratory Panel 2.1 on nasal swabs. No rebound patients required additional treatment or hospitalization. Median C-reactive protein (CRP) level was lower at rebound than during acute COVID-19, whereas neutrophil and lymphocyte counts and SARS-CoV-2 PCR cycle thresholds (Ct) were similar across groups (Fig-1A-D) with low or undetectable serum nucleocapsid antigen levels at rebound (Fig-1E). Infectious replication-competent SARS-CoV-2 was isolated from the nasal swab of 3/4 controls and 1/7-rebound patients (Patient-1). No mutations associated with NMV resistance were identified. Inflammatory markers associated with myeloid cell activation and severe COVID-19,^10^ such as IL-6 or IL-8, were downtrending at rebound, whereas markers of T-cell activation like interferon-gamma and soluble CD25 were stable or increasing (Fig-S1).

**Figure-1:**
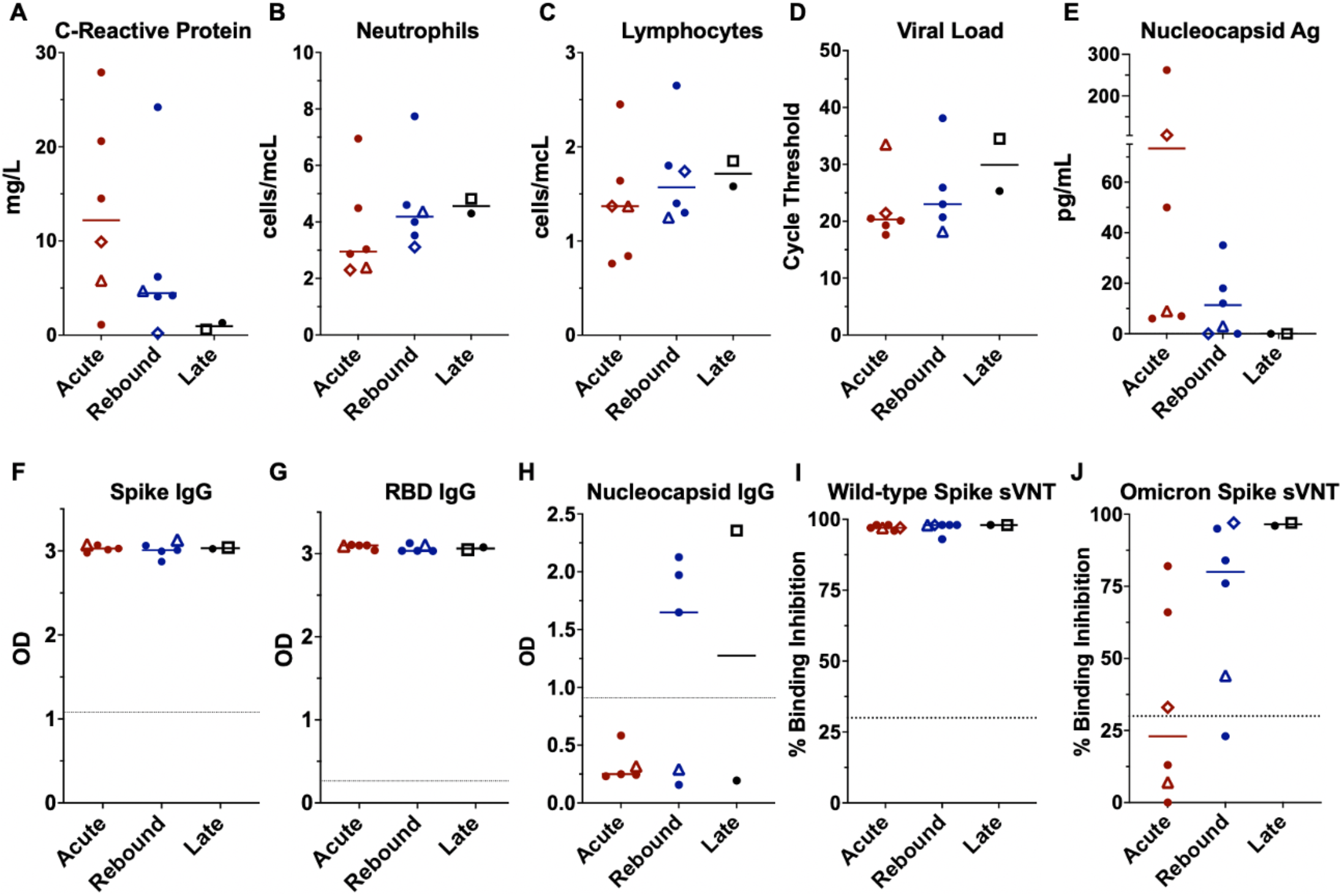
Comparison of clinical laboratories, virologic measurements, and antibody responses across the groups. Lines represent median and points represent individual results. The two longitudinal patients are identified by an open diamond (patient 1) and open triangle (patient 2), respectively. The open square represents the COVID-19 rebound patient that did not receive NMV-r. Clinical values for C-reactive protein, absolute neutrophil count, and absolute lymphocyte count across the acute (red), rebound (blue), and late presenting (black) COVID-19 cohorts (A, B, C). SARS-CoV-2 cycle threshold from nasal swab samples (D) and serum nucleocapsid antigen (E). Antibody levels by enzyme-linked immunosorbent assay (ELISA) against the spike protein, spike - receptor binding domain (RBD), and the nucleocapsid protein (F, G, H) presented as optical density (OD). ELISA data not available for longitudinal patient-1 (diamond). Surrogate viral neutralization test (sVNT) to detect neutralizing antibodies against the wild-type (I) and Omicron (J) spike protein presented as percent binding inhibition. Dotted lines represent the cut-off for a positive result for the antibody tests (F-J). Abbreviations: Ag – antigen, OD – optical density, RBD – receptor-binding domain, sVNT – surrogate viral neutralization test.

Anti-S and anti-RBD IgG antibodies were at high levels in both groups (Fig-1F-G), consistent with prior vaccination. Anti-nucleocapsid IgG antibodies were absent in acute and detectable in 4 rebound patients including the untreated patient (Fig-1H). IgM and IgA responses were variable (Fig-S2A-E). Surrogate VNTs^9^ assays showed high levels of wild-type spike neutralizing antibodies in all patients with lower percent binding inhibition to Omicron-spike protein, especially in the acute group (Fig-1I-J). More rebound patients had detectable neutralizing antibodies against Omicron-spike that inversely correlated with serum antigen (Fig-S2F).

CD4 and CD8 T-cell counts increased in rebound and late cases (Fig-2A-B). Robust T-cell responses against the spike protein with more antigen-specific, cytokine-producing (IFNγ, TNFα) CD4^+^ T-cells were seen in those with rebound or late presentation (Fig-2C-E). CD4^+^ T-cell responses against nucleocapsid and membrane proteins were also higher in the late and rebound cases (Fig-2C-E). Two patients were evaluated longitudinally (Table-S3). They both exhibited significant drops in CRP and serum nucleocapsid antigen (Fig-2F-G) at rebound, with concomitant increases in neutralizing antibodies and SARS-CoV-2 specific CD4^+^ T-cells (Fig-2H-I).

**Figure-2:**
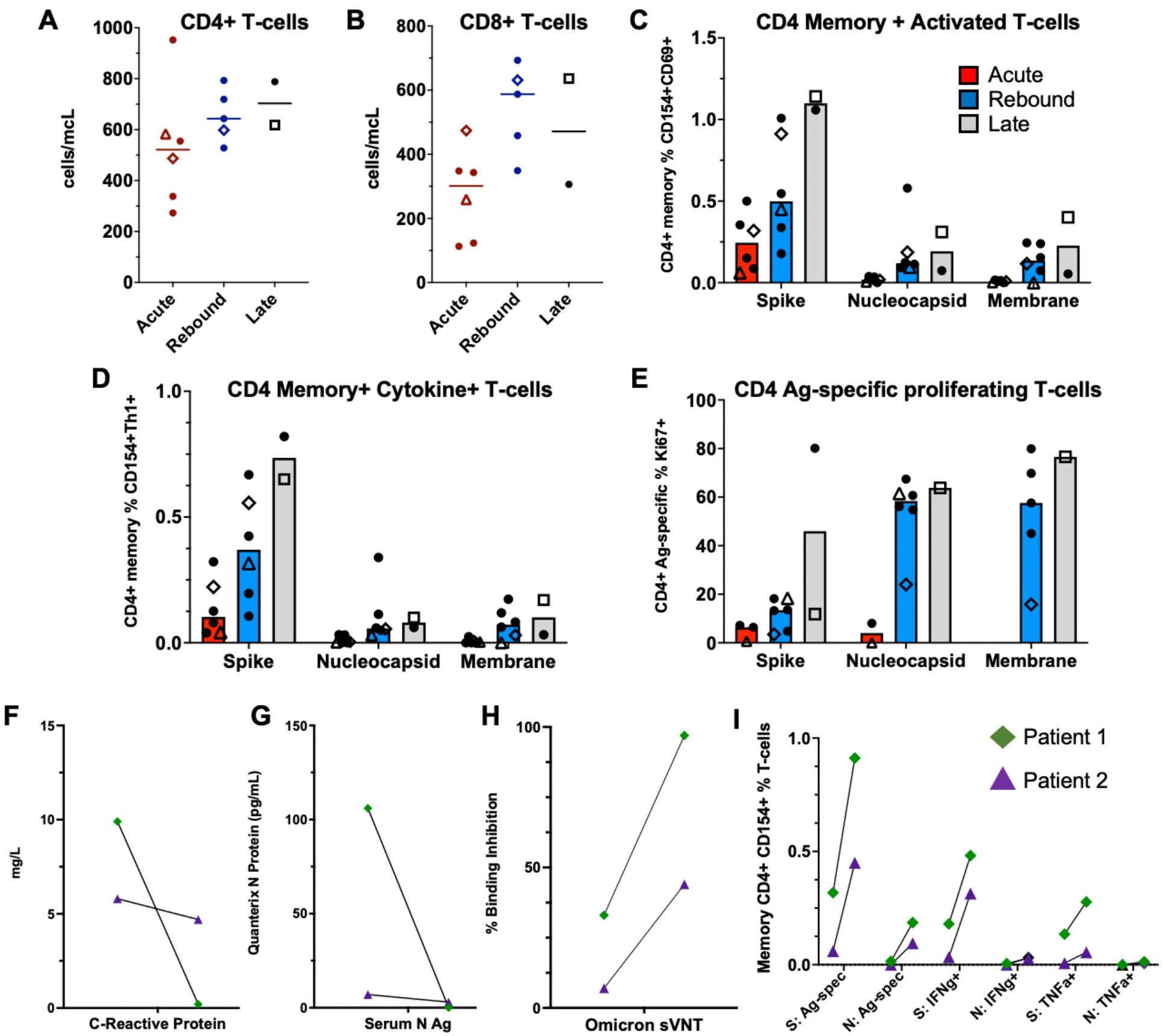
Comparison of T-cell responses across the groups and longitudinal immune responses of the two patients with sampling at acute and rebound timepoints. Absolute T-cell counts compared across groups (A-B). Lines represent median values and points represent individual results. The two-longitudinal patients are identified by an open triangle and open diamond. The empty square represents the COVID-19 rebound patient that did not receive NMV-r. T-cell counts not available for longitudinal patient-2 (triangle) at rebound timepoint. T-cell stimulations were performed with peptide pools corresponding to spike, nucleocapsid, and membrane proteins as listed on the x-axis. Bars represent medians and groups are defined as acute (red), rebound (blue), and late presentations (gray). SARS-CoV-2 specific CD4 T-cell responses are highlighted by memory, cytokine-producing (CD154+IFNγ+, CD154+TNFα+ or CD154+IL-2+), activated (CD154+CD69+), or antigen-specific proliferating T-cells (Ki67+) (C-E). For phenotyping of Ki67+ cells, a threshold of at least 20-events and a 2-fold increase over unstimulated cells was used, and samples were excluded if they did not meet these thresholds (E). Serum nucleocapsid antigen (Ag) and C-reactive protein trends from the two longitudinal patients (F-G). T-cell responses and neutralizing antibodies from the acute and rebound presentation for two patients with longitudinal samples (H-I). T-cell responses are from spike (S) and nucleocapsid (N) stimulations. Antigen-specific CD4 T-cells defined by (CD154+CD69+, CD154+IFNγ+ and CD154+TNFα+), and neutralizing antibodies represented by percent binding inhibition on the surrogate virus neutralization test (sVNT). Abbreviations: Ag – antigen, S – spike, N – nucleocapsid, Ag-spec – antigen-specific, sVNT - surrogate virus neutralization test, N Ag – nucleocapsid antigen.

## Discussion

Nirmatrelvir/ritonavir has been a long-awaited addition in the COVID-19 therapeutic armamentarium as an outpatient oral medication that can improve disease prognosis in high-risk patients. Cases of clinical rebound after NMV-r reported recently have raised concerns about clinical deterioration and interference of early antiviral administration with adaptive immune responses. The licensing trial did not identify significant differences in viral rebound incidence among NMV-r versus placebo in unvaccinated patients during the delta variant wave.^2^ Currently, NMV-r is widely used in breakthrough infections by the Omicron variant, which may impact the incidence of clinical rebound. Larger studies are required to determine the incidence and risk factors for rebound COVID-19 in those treated versus not treated with NMV-r.

In our case series, which includes one patient with rebound without prior antiviral therapy, no patients developed severe disease or required additional therapy. High levels of SARS-CoV-2 anti-S IgG antibodies were detected in all patients consistent with prior vaccination. Anti-nucleocapsid IgG and Omicron-specific neutralizing antibodies were higher in patients with rebound consistent with development of humoral immunity, that usually occurs 2–3-weeks post-infection.^11^ Additionally, we detected rising T-cells and robust SARS-CoV-2 specific T-cell responses at rebound, that were greater than those in early acute COVID-19. Two patients with longitudinal sampling demonstrated an increase in both antibody and cellular immune responses during rebound compared to their acute presentation. These findings refute the hypothesis that impaired adaptive immune responses contribute to symptomatic rebound.

Resistance mutations were not identified at COVID-19 rebound, consistent with prior reports.^3,12^ SARS-CoV-2 was isolated from culture in one of seven patients with rebound and in three of four tested controls with acute infection probably signifying higher transmission potential in early disease. In a recent report, virus growth was observed in samples from three of seven rebound patients after NMV-r^12^, however, different methodologies may explain this disparity. Interestingly, our rebound patient with culturable virus had underlying immune suppression raising the question of longer treatment for immunocompromised persons.

Overall, our findings of lower levels of serum nucleocapsid antigen and downtrending innate immune markers with an emerging adaptive immune response in rebound COVID-19, do not support uncontrolled viral replication driving inflammation with significant risk for impending disease progression. Increases in total and virus-specific T-lymphocytes, biomarkers of T-cell activation, and antibodies suggest that the rebound symptoms may in fact be partially driven by the emerging immune response against residual viral antigens possibly shed from dying infected cells due to cytotoxicity and tissue repair throughout the respiratory tract.^13^ Symptoms may be more clinically evident after use of potent antiviral treatment with quick clinical improvement and re-appearance of antigen at the time of a maturing immune response.

In conclusion, this case series provides important insights into the pathophysiology of rebound COVID-19. None of these patients developed severe disease, and adaptive immunity against SARS-CoV-2 appeared intact. Our findings support the need for isolation and the consideration for prolonged or additional therapies for immunocompromised patients who cannot rely on adaptive immune responses. Evaluation of larger cohorts is required to further assess the incidence, clinical and, importantly, epidemiologic implications of rebound COVID-19.

All authors declare no conflicts of interest.

## Data Availability

All data produced in the present work are contained in the manuscript

## Ethics Approval

All patients and healthy controls evaluated at the NIH provided written informed consent and were enrolled on NCT04401436 or NCT00001281. Both of these protocols were approved by the National Institutes of Health Institutional Review Board.

## Acknowledgements

Funding for this study was provided in part by the Division of Intramural Research, National Institute of Allergy and Infectious Diseases, National Institutes of Health. The content of this publication does not necessarily reflect the views or policies of the Department of Health and Human Services, nor does the mention of trade names, commercial products, or organizations imply endorsement by the U.S. Government.

## Notes

### Competing Interest Statement

The authors have declared no competing interest.

### Author Declarations

All patients and healthy controls were evaluated at the National Institutes of Health and provided written informed consent and were enrolled on the "COVID-19 Associated Lymphopenia Pathogenesis Study in Blood" or a separate protocol for healthy volunteers (NCT00001281) both of which were approved by the National Institutes of Health Institutional Review Board

## References

1. Emergency Use Authorization (EUA) for Paxlovid (nirmatrelvir tablets co-packaged with ritonavir tablets) Center for Drug Evaluation and Research (CDER) Review. In: Center for Drug Evaluation and Research (CDER); 2021.

2. Hammond J, Leister-Tebbe H, Gardner A, et al. Oral Nirmatrelvir for High-Risk, Nonhospitalized Adults with Covid-19. N Engl J Med. 2022;386(15):1397–1408.

3. Charness M, Gupta K, Stack G, et al. Rapid Relapse of Symptomatic Omicron SARS-CoV-2 Infection Following Early Suppression with Nirmatrelvir/Ritonavir. medRxiv Preprint. 2022.

4. Gupta K, Strymish J, Stack G, Charness M. Rapid Relapse of Symptomatic SARS-CoV-2 Infection Following Early Suppression with Nirmatrelvir/Ritonavir. medRxiv Preprint. 2022.

5. Bennett RS, Postnikova EN, Liang J, et al. Scalable, Micro-Neutralization Assay for Assessment of SARS-CoV-2 (COVID-19) Virus-Neutralizing Antibodies in Human Clinical Samples. Viruses. 2021;13(5).

6. Nussenblatt V, Roder AE, Das S, et al. Yearlong COVID-19 Infection Reveals Within-Host Evolution of SARS-CoV-2 in a Patient With B-Cell Depletion. J Infect Dis. 2022;225(7):1118–1123.

7. Group A-TBS, Lundgren JD, Grund B, et al. Responses to a Neutralizing Monoclonal Antibody for Hospitalized Patients With COVID-19 According to Baseline Antibody and Antigen Levels : A Randomized Controlled Trial. Ann Intern Med. 2022;175(2):234–243.

8. Klumpp-Thomas C, Kalish H, Drew M, et al. Standardization of ELISA protocols for serosurveys of the SARS-CoV-2 pandemic using clinical and at-home blood sampling. Nat Commun. 2021;12(1):113.

9. Tan CW, Chia WN, Qin X, et al. A SARS-CoV-2 surrogate virus neutralization test based on antibody-mediated blockage of ACE2-spike protein-protein interaction. Nat Biotechnol. 2020;38(9):1073–1078.

10. Lucas C, Wong P, Klein J, et al. Longitudinal analyses reveal immunological misfiring in severe COVID-19. Nature. 2020;584(7821):463–469.

11. Long QX, Liu BZ, Deng HJ, et al. Antibody responses to SARS-CoV-2 in patients with COVID-19. Nat Med. 2020;26(6):845–848.

12. Boucau J, Uddin R, Marino C, et al. Virologic characterization of symptom rebound following nirmatrelvir-ritonavir treatment for COVID-19. medRxiv preprint. 2022.

13. Sefik E, Qu R, Junqueira C, et al. Inflammasome activation in infected macrophages drives COVID-19 pathology. Nature. 2022.

