## Supplement for "COVID-19 redux: clinical, virologic, and immunologic evaluation of clinical rebound after nirmatrelvir/ritonavir"

### Online Supplement

#### Supplemental Methods

##### Clinical Protocol

All participants were evaluated at the National Institutes of Health (NIH) between December 2021 and May 2022 and enrolled in the clinical protocol: COVID-19–associated Lymphopenia Pathogenesis Study in Blood (CALYPSO), NCT04401436. In brief, this protocol evaluates adults (18-years or older) with a diagnosis of COVID-19 by a molecular or other commercial assay, or people who have recovered from COVID-19. Participation includes clinical evaluation, standard of care clinical management as indicated, laboratory studies, and research blood collection for storage of peripheral blood mononuclear cells (PBMCs), serum, and plasma. Additionally, plasma samples were obtained from seven healthy volunteers on a separate NIH IRB-approved protocol (NCT00001281). This was a convenience sample to provide a healthy control comparison for the biomarker analysis and consisted of 5-women and 2-men with a median age of 59-years (range: 45-66 years). All participants in both protocols provided written informed consent prior to any study procedures in accordance with the Declaration of Helsinki.

##### Viral Sequencing

Initial detection and estimation of viral load by cycle threshold (Ct values) was performed using Hologic® Panther Fusion® SARS-CoV-2 assay. Viral RNA sequencing was performed on SARS-CoV-2 positive nasal swab samples collected from patients with rebound symptoms as previously described (1). Amplification of the viral genome was performed using a modified version of the ARTIC consortium protocol for nCoV-2019 sequencing (https://artic.network/ncov-2019) and the methods available at https://github.com/GhedinSGS/SARS-CoV-2_analysis. All libraries were prepped using the Nextera XT library preparation kit (Nextera), scaled down to 0.25x of the manufacturer's instructions and libraries were sequenced on the Illumina MiSeq using the 2x300 bp paired end protocol. Following sequencing, samples were processed using the snakemake pipeline available at https://github.com/GhedinSGS/SARS-CoV-2_analysis. The output from an in-house variant calling pipeline, timo, was used to identify consensus changes. All analyses performed for this study are available at https://github.com/GhedinSGS/paxlovid-study.

**Viral Culture**

Viral cultures were also performed on each sample by adding them to individual flasks containing Vero E6 cells expressing the transmembrane serine protease, TMPRSS2 (VeroE6/TMPRSS2), obtained from the Japanese Cell Culture collection (Osaka, Japan). Inoculations were performed as previously described (2). The infected cells were incubated at 37º C with 5% CO2 and observed daily for cytopathic effects (CPE). Upon observation of CPE the virus supernatant is harvested and sequenced. Finally, SARS-CoV-2 viral nucleocapsid protein levels were measured in serum using a single-molecule immune bead assay (Quanterix, Billerica, MA) (3).

**Serologic Assays**

SARS-CoV-2 anti-spike (anti-S) and anti-receptor binding domain (anti-RBD) antibody responses were analyzed by enzyme-linked immunosorbent assay (ELISA) using a previously validated technique (4). In this study, a threshold of three standard deviations above the mean of the negative controls for both the spike and RBD was determined to have a specificity of 100% for a positive result, and therefore was considered the threshold for positivity. Levels of IgG, IgM, and IgA were determined for anti-S and anti-RBD and compared between the rebound cases and controls. Anti-nucleocapsid antibodies were evaluated only for IgG and IgM.

The GenScript cPass assay, a surrogate viral neutralization test (sVNT) which has been shown to detect neutralizing antibodies (5), was also performed to evaluate for the presence of anti-S neutralizing antibodies against wild-type SARS-CoV-2 and the Omicron variant. This assay measures anti-S antibody-mediated inhibition of the interaction between the SARS-CoV-2 spike-RBD and the angiotensin-converting enzyme-2 (ACE2) receptor. The result is reported as percent binding inhibition of this interaction. Diluted serum samples (1:10) were mixed with equal volume of HRP conjugated RBD antigens and incubated for 30 minutes at 37º C. Then 100 ul of conjugates were transferred to the microplate coated with ACE2-receptors and incubated for 15-minutes at 37º C. The microplate was washed 4-times and incubated with TMB substrate in the dark for 15-minutes at room temperature for color development. Absorbance was measured at 450 nm after addition of stop solution. The results were calculated by the formula: Inhibition (%) = (1- OD value of sample/OD value of negative control) x 100. The results were interpreted as positive if ≥ 30% inhibition. For detection of neutralizing antibodies to the BA.1 Omicron variant, the wild type S-RBD was replaced by the BA.1 S-RBD in the assay.

**T-cell stimulation assays**

Frozen PBMCs were thawed into pre-warmed R10 media (RPMI 1640, 10% FBS, 2 mM L-glutamine, 100 U/ml penicillin, and 100 µg/ml streptomycin) containing DNase I and rested for 1 hour (37ºC/5% $CO_2$). $1x10^6$ to $1.5x10^6$ PBMCs were aliquoted into 96-well V-bottom plates in 200 µL R10 and stimulated with peptide pools (15mers overlapping by 11 amino acids) corresponding to the SARS-CoV-2 spike (wild-type), nucleocapsid and membrane proteins (2µg/mL final concentration; Genscript). Protein transport inhibitors (ThermoFisher; cat. 00-4980-03) were included during the stimulation. Following stimulation for 6 hours (37ºC/5% $CO_2$), PBMCs were washed with PBS, incubated with LIVE/DEAD Fixable Blue Dead Cell Stain for 10 minutes and subsequently stained for surface markers (anti-CD3, anti-CD4, anti-CD8, anti-TCR$\gamma\delta$, anti-CCR7, anti-CD45RA and anti-PD-1) for 20 minutes. All incubation steps during the staining were performed at room temperature. Cells were washed in PBS +

2% FBS, and subsequently fixed for 30 minutes (FoxP3/Transcription Factor Staining Buffer Set; ThermoFisher). Fixed PBMCs were washed in Permeabilization buffer and stained for intracellular proteins (anti-CD69, anti-CD154, anti-IFNγ, anti-IL-2, anti-IL-4, anti-IL-13, anti-IL-21, anti-TNFα, anti-IL17 and anti-Ki67) for 30 minutes. Cells were again washed with Permeabilization buffer and fixed with 1% paraformaldehyde. Data were acquired on a modified BD FACSymphony and analyzed using FlowJo software (version 10.7.1). Cytokine frequencies were background subtracted using a DMSO only condition. Any CD4+ Th1-type response was determined using Boolean gating to include CD154+IFNγ+, CD154+IL-2+ or CD154+TNFα+ responses. Two peptide pools were utilized to measure Spike-specific responses, Spike pool A (peptides 1-160; residues 1-651) and Spike pool B (peptides 161-316; residues 641-1273) and responses from the two pools were summed to determine the response to full-length Spike.

**Antibodies for T cell stimulation assay**

The following antibodies were purchased from BD Biosciences: CD3 BV786 (Clone SK7; cat. 563800), CD4 BUV496 (Clone SK3; cat. 612936), CD8 BUV805(Clone SK1; cat. 612889), IFNγ BB700 (Clone B27; cat. 566394), TNFα BV750 (Clone MAb11; cat. 566359), IL-21 AlexaFluor647 (Clone 3A3-N2.1; cat. 560493), IL-13 PE (Clone JES10-5A2; cat. 554571) Ki67 BV650 (Clone B56; cat. 563757) and CCR7 BUV395 (Clone 150503; custom conjugation). Antibodies from Biolegend included: IL-2 BV421 (Clone MQ1-17H12, cat. 500328), CD154 BV605 (Clone 24-31; cat. 310826), CD69 BV711 (Clone FN50, cat. 310944), IL-4 PE (Clone MP4-25D2, cat. 500810), CD45RA BV570 (Clone HI100; cat. 304132) and PD-1 PE-Cy7 (Clone EH12.2H7; cat. 329918). Anti-TCR$\gamma\delta$ APC-Vio770 (Clone REA591, cat. 130-113-509) was from Miltenyi.

**Soluble Biomarkers**

Plasma samples were used to measure the following biomarkers according to the manufacturer's instructions. Plasma levels of interleukin (IL)-6, IL-8, IL-10, IL-18, tumor necrosis factor-α (TNFα), interferon-γ (IFNγ), and chemokine (C-X-C motif) ligand 10 (CXCL10) were quantified by electrochemiluminescence assays from Meso Scale Discovery platforms (Gaithersburg, MD, USA). Plasma levels of CXCL-9, soluble CD25 (sCD25), IL-18 binding protein (BP), and sCD14 were quantified using enzyme-linked immunoassay (ELISA) kits from R&D Systems (Minneapolis, MN, USA).

**Statistical Analysis**

All statistical analyses were conducted using GraphPad Prism (version 9) and R software (version 4.1.0).

1. Nussenblatt V, Roder AE, Das S, de Wit E, Youn JH, Banakis S, et al. Yearlong COVID-19 Infection Reveals Within-Host Evolution of SARS-CoV-2 in a Patient With B-Cell Depletion. J Infect Dis. 2022;225(7):1118-23.

2. Bennett RS, Postnikova EN, Liang J, Gross R, Mazur S, Dixit S, et al. Scalable, Micro-Neutralization Assay for Assessment of SARS-CoV-2 (COVID-19) Virus-Neutralizing Antibodies in Human Clinical Samples. Viruses. 2021;13(5).

3. Group A-TBS, Lundgren JD, Grund B, Barkauskas CE, Holland TL, Gottlieb RL, et al. Responses to a Neutralizing Monoclonal Antibody for Hospitalized Patients With COVID-19 According to Baseline Antibody and Antigen Levels : A Randomized Controlled Trial. Ann Intern Med. 2022;175(2):234-43.

4. Klumpp-Thomas C, Kalish H, Drew M, Hunsberger S, Snead K, Fay MP, et al. Standardization of ELISA protocols for serosurveys of the SARS-CoV-2 pandemic using clinical and at-home blood sampling. Nat Commun. 2021;12(1):113.

5. Tan CW, Chia WN, Qin X, Liu P, Chen MI, Tiu C, et al. A SARS-CoV-2 surrogate virus neutralization test based on antibody-mediated blockage of ACE2-spike protein-protein interaction. Nat Biotechnol. 2020;38(9):1073-8.

Supplemental Table 1: Laboratory Values

| | **Acute (N=6)** | **Rebound after NMV-r (N=6)** | **Late (N=2)** |
|---|---|---|---|
| **Days from initial symptom onset to sample collection, median [Min, Max]** | 3 [2-4] | 16 [11-17] | 11 [8-14] |
| **Absolute neutrophil count, median [Min-Max] (cells/µL)** | 2950 [2300-6950] | 4190 [3110-7740] | 4560 [4300-4820] |
| **Absolute lymphocyte count, median [Min-Max] (cells/µL)** | 1370 [760-2450] | 1570 [1250-2650] | 1720 [1580-1850] |
| **C-reactive protein, median [Min-Max] (mg/L)** | 12.2 [1.1-27.9] | 4.45 [0.2-24.2] | 0.95 [0.6-1.3] |
| **Ferritin, median [Min-Max] (µg/L)** | 178 [26-551] | 192 [30-251] | 72.5 [36-109] |
| **CD4 T cells, median [Min-Max] (cells/µL)** | 521 [273-952] | 643 [528-793] | 703 [618-788] |
| **CD8 T cells, median [Min-Max] (cells/µL)** | 301 [113-474] | 587 [349-693] | 471 [306-636] |
| **LDH, median [Min-Max] (units/L)** | 171 [100-229] | 182 [138-190] | 164 [123-205] |
| **D-dimer, median [Min-Max] (µg/mL)** | 0.37 [0.27-2.22] | 0.36 [0.27-0.50] | 0.50 [0.33-0.66] |

Supplemental Table 2: Individual Patient Clinical Courses and Laboratory Values During Rebound

| | NMV-r Recipient Rebound Patients | | | | | | NMV-r Nonrecipient Rebound |
|---|---|---|---|---|---|---|---|
| | 1 | 2 | 3 | 4 | 5 | 6 | |
| **Initial symptoms** | Upper respiratory + constitutional | Upper and lower respiratory | Upper respiratory | Upper respiratory | Upper respiratory + constitutional | Upper respiratory + constitutional | Upper respiratory + constitutional |
| **Day of illness NMV-r started** | 3 | 2 | 1 | 3 | 1 | 4 | NA |
| **Day of illness symptoms returned** | 15[a] | 11[a] | 11[b] | 14 | 14 | 11[a] | 9 |
| **Day symptoms returned after completing NMV-r** | 8 | 5 | 6 | 7 | 9 | 3 | NA |
| **Days from initial symptom onset to visit** | 16 | 11 | 13 | 16 | 16 | 17 | 14 |
| **Rebound symptom severity** | Milder[c] | Milder[c] | Worse[d] | Milder[c] | Similar[e] | Milder[c] | Milder[c] |
| **Temperature during evaluation (°Celsius)** | 36.6 | 37.4 | 36.5 | 36.4 | 36.8 | 36.4 | 36.5 |
| **Initial heart rate during evaluation (beats/minute)** | 86 | 74 | 120 | 93 | 60 | 77 | 70 |
| **Oxygen saturation during evaluation (percent)** | 97 | 99 | 98 | 97[f] | 97 | 99 | 100 |
| **Absolute neutrophil count (cells/µL)** | 3110 | 4370 | 4600 | 4000 | 7740 | 3520 | 4300 |
| **Absolute lymphocyte count (cells/µL)** | 1740 | 1250 | 2650 | 1800 | 1400 | 1300 | 1850 |
| **C-reactive protein (mg/L)** | 0.2 | 4.7 | 24.2 | 4.1 | 6.2 | 4.2 | 0.6 |
| **Ferritin (µg/L)** | 251 | 30 | 226 | 151 | 228 | 157 | 109 |

| | | | | | | | |
|---|---|---|---|---|---|---|---|
| **CD4 T cells (cells/µL)** | 598 | NA | 793 | 719 | 643 | 528 | 618 |
| **CD8 T cells (cells/µL)** | 631 | NA | 693 | 587 | 349 | 458 | 636 |
| **LDH (units/L)** | 183 | 138 | 172 | 184 | 190 | 180 | 123 |
| **D-dimer (µg/mL)** | 0.27 | 0.36 | 0.40 | 0.36 | 0.50 | 0.36 | 0.66 |

[a] Repeat home rapid antigen turned negative, then positive again during rebound.

[b] Repeat home rapid antigen turned weakly positive, then became strongly positive again during rebound.

[c] Typical upper respiratory symptoms returned, no other symptoms.

[d] Upper respiratory symptoms returned and developed new lower respiratory symptoms (dyspnea on exertion).

[e] Upper respiratory symptoms as well as constitutional symptoms (fevers and myalgias) returned.

[f] Oxygen saturation was 94% after exertion and returned to normal range with rest.

Supplemental Table 3: Patient 1 and 2 Acute and Rebound Timepoints

| | Patient 1 Acute | Patient 1 Rebound | Patient 2 Acute[a] | Patient 2 Rebound |
|---|---|---|---|---|
| **Days from initial symptom onset to sample** | 3 | 16 | 4 | 11 |
| **Absolute neutrophil count (cells/µL)** | 2300 | 3110 | 2390 | 4370 |
| **Absolute lymphocyte count, median (cells/µL)** | 1370 | 1740 | 1370 | 1250 |
| **C-reactive protein (mg/L)** | 9.90 | 0.20 | 5.80 | 4.70 |
| **Ferritin (µg/L)** | 216 | 251 | 38.0 | 30.0 |
| **CD4 T cells (cells/µL)** | 487 | 598 | 583 | NA |
| **CD8 T cells (cells/µL)** | 474 | 631 | 259 | NA |
| **D-dimer (µg/mL)** | 0.27 | 0.27 | 0.37 | 0.36 |
| **LDH (units/L)** | 147 | 183 | 194 | 138 |

[a] Baseline evaluations were done after first 24 hours of NMV/r.

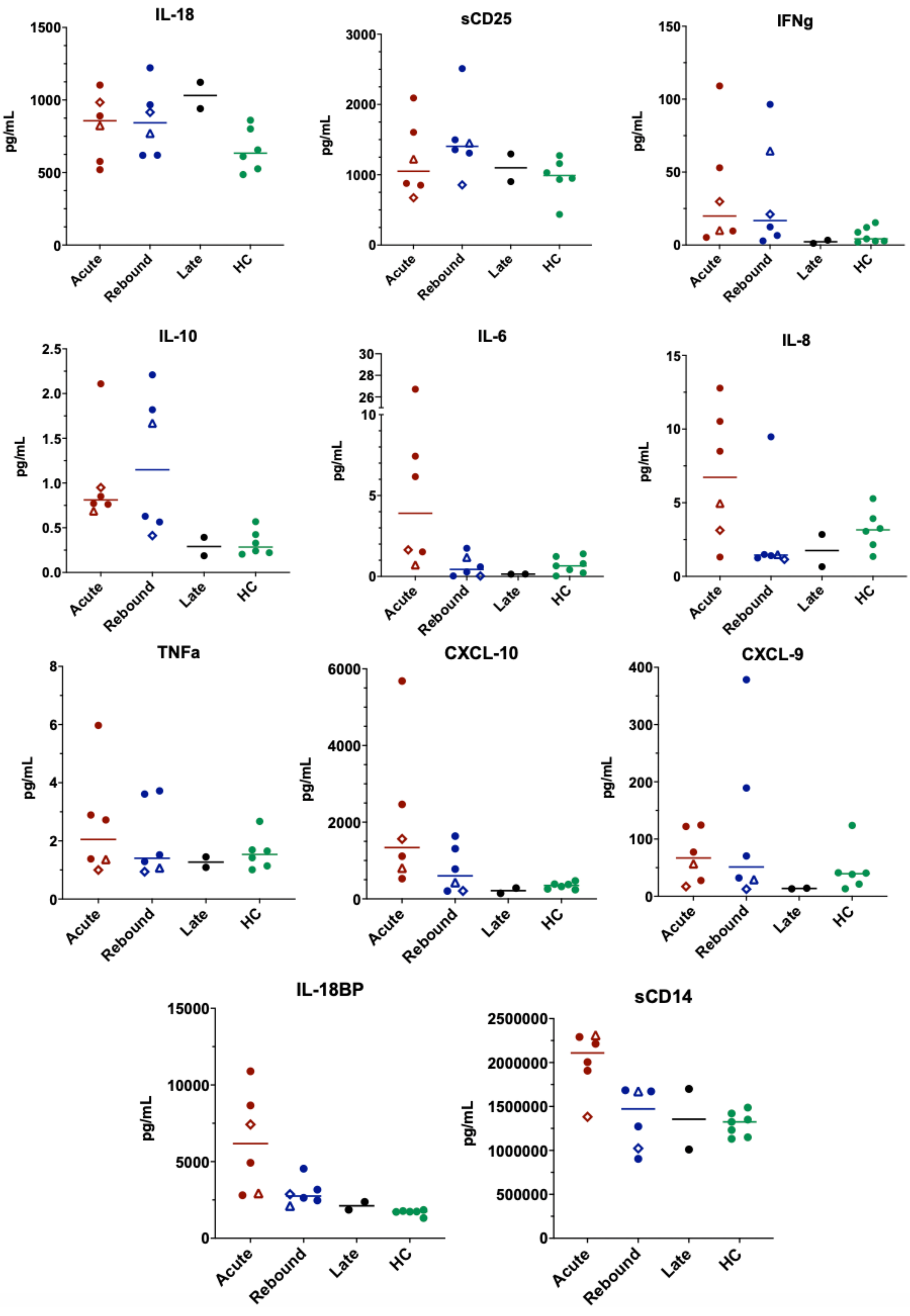
IL-18
sCD25
IFNg
IL-10
IL-6
IL-8
TNFa
CXCL-10
CXCL-9
IL-18BP
sCD14
pg/mL
Acute
Rebound
Late
HC

**Supplemental Figure-1: Innate and adaptive biomarkers across study groups compared to healthy controls.** Lines represent medians and points represent individual results across the acute (red), rebound (blue) and late presentation (black) COVID-19 clinical groups compared to healthy control population (green). Healthy controls consisted of 5-women and 2-men with a median age of 59-years (range: 45-66 years). These samples were unmatched and included to provide a baseline range for these biomarkers in an otherwise healthy population. The two longitudinal COVID-19 rebound patients are identified by an open diamond (Patient-1) and open triangle (Patient-2). The open square represents the COVID-19 rebound patient that did not receive NMV-r. Innate biomarkers (IL-6, IL-8, TNFα, CXCL-10, sCD14, and IL-18BP) classically increased in acute COVID-19 are down trending at time of rebound. Adaptive biomarkers representing T-cell activation (IFNγ, CXCL-9, sCD25) were stable or increasing at rebound consistent with a developing T-cell response. Abbreviations: IL – interleukin, sCD25 – soluble CD25, IFNγ – interferon-γ, TNFα – tumor necrosis factor-α, CXCL-10 - chemokine (C-X-C motif) ligand 10, IL-18BP – interleukin-18 binding protein.

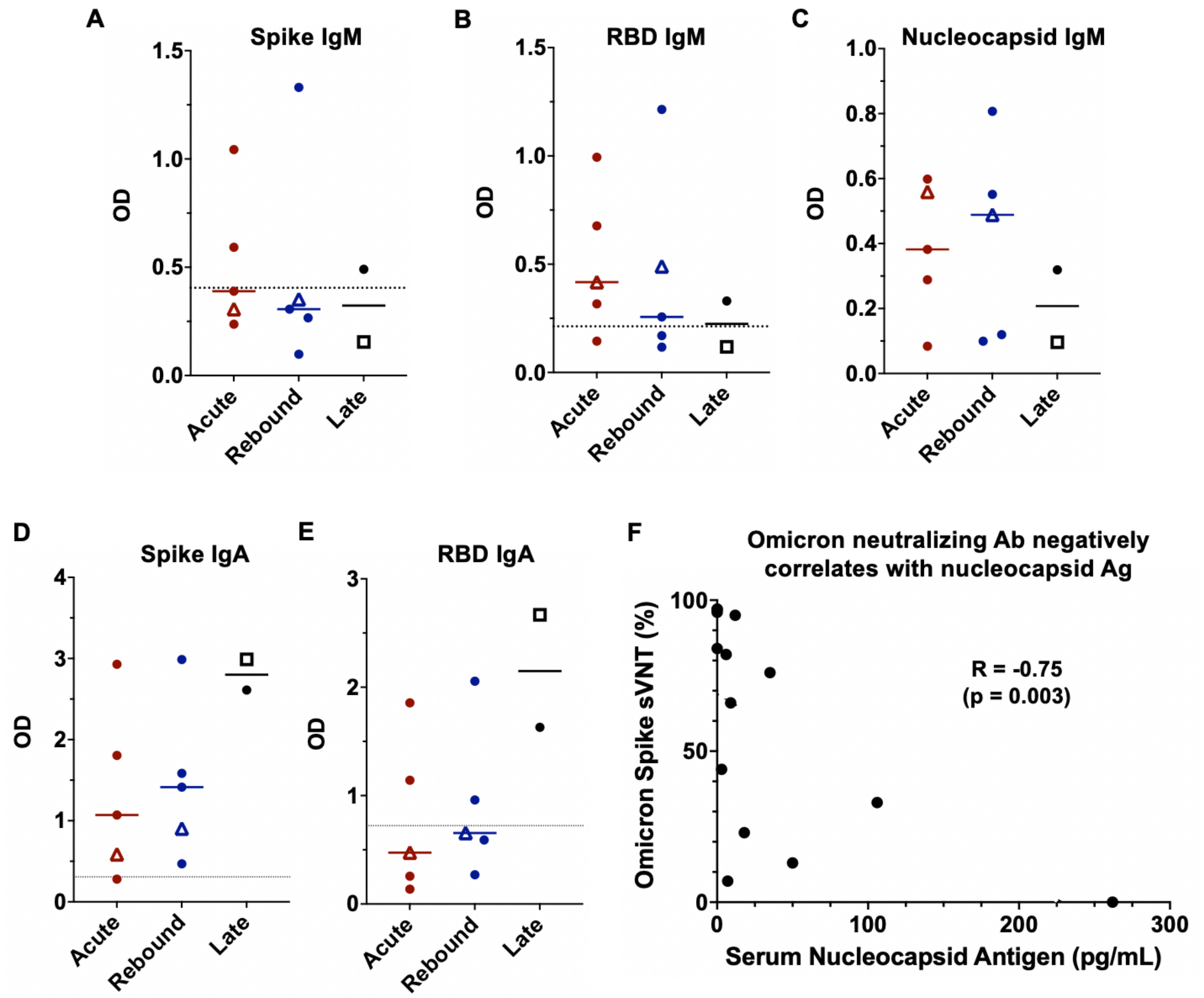


**Supplemental Figure-2: Summary of IgM and IgA antibody responses against SARS-CoV-2 proteins across the three groups.** Lines represent medians and points represent individual results. Longitudinal patient-2 is represented by an open triangle. ELISA data not available for longitudinal patient-1 (diamond). The open square represents the COVID-19 rebound patient that did not receive NMV-r. IgM and IgA antibody responses against spike protein and receptor binding domain (A-B, D-E). IgM anti-nucleocapsid (N) antibody response (C). Surrogate virus neutralization test (sVNT) presented as percent binding inhibition against Omicron SARS-CoV-2 spike protein negatively correlated with serum nucleocapsid antigen (pg/mL) (R = -0.30 by Spearman's correlation with p = 0.028) (F). Abbreviations: OD – optical density, RBD – receptor binding domain, N – nucleocapsid protein, sVNT – surrogate virus neutralization test, Ab – antibody, Ag – antigen.
